# Cross-Cultural Adaptation and Validation Study of the Brazilian Version of the Multidimensional State Boredom Scale (MSBS)

**DOI:** 10.1101/2025.05.31.25328648

**Authors:** Tiago Figueiredo

## Abstract

This study aimed to conduct a transcultural adaptation of the Multidimensional State of Boredom Scale (MSBS) for the Brazilian sociocultural context. A total of 329 volunteers (79% female and 21% male), aged between 12 and 68 years (M = 33.08, SD = 12.44), participated in the study. The process involved expert back-translation carried out by bilingual language professionals and a boredom specialist. Participants completed the MSBS and a socioeconomic questionnaire. A committee of judges evaluated the content validity, indicating excellent levels. Confirmatory factor analysis (CFA) revealed that the scale retained its multidimensional structure, consisting of five factors (Disengagement, High Arousal, Inattention, Low Arousal, and Time Perception subscales), consistent with the original version of the MSBS. Additionally, the Brazilian version of the MSBS demonstrated strong reliability. These findings suggest that the MSBS has excellent psychometric properties in the Brazilian context. The study contributes to a deeper understanding of the state of boredom, its association with mental disorders, and facilitates cross-cultural comparisons within the general population.

## Introduction

There are several definitions of boredom across various fields of study (Eastwood et al., 2012). In psychology, efforts to define boredom as a mental state intrinsic to human experience date back to the early 20th century. In 1903, Lipps characterized boredom as a psychological state marked by negative emotions—specifically, an emotional state of displeasure—resulting from diminished psychic stimulation due to environmental factors. Since this initial definition, the concept of boredom has been further developed through contributions from diverse theoretical perspectives. In 1993, Mikulas and Vodanovich proposed an integrated definition of boredom, describing it as a mental state characterized by low excitability and a sense of displeasure, often stemming from a lack of stimulation in one’s environment. Nearly two decades later, in 2012, John Eastwood and his collaborators introduced a widely referenced definition of boredom in contemporary academic literature. They defined boredom as an aversive and unpleasant mental state that involves a diminished desire and an inability to engage in fulfilling activities. Thus, boredom is an aversive mental state that arises from unpleasant contexts and cognitive disengagement (Gerritsen et al., 2014; Danckert & Elpidorou, 2023).

In the field of clinical psychiatry research, boredom has been identified as a factor with a bidirectional role in relation to psychopathology. On one hand, chronic boredom is frequently associated with negative mood and depressive symptoms, serving as a driving force that can lead to adverse consequences such as poor academic performance, overeating, alcohol consumption, and delinquency in individuals across various developmental stages (Jarvis & Seifert, 2002; Lee & Zeldeman, 2019; Panda et al., 2021; Biocalti et al., 2016; Spaeth et al., 2015). On the other hand, several internal psychological factors have been linked to boredom proneness. These include cognitive factors, such as poor attention abilities and executive dysfunction (Hunter & Eastwood, 2018); motivational factors, which encompass extrinsic motivation (i.e., engaging in activities for external rewards) and a strong desire to minimize pain or maximize pleasure (Mercer-Lynn, Hunter, & Eastwood, 2013); volitional or self-regulatory factors, which involve poor self-control (Isacescu & Danckert, 2018), a preference for “doing the right thing” rather than taking action (Mugon et al., 2018), and a state rather than action orientation (i.e., the tendency to focus on one’s thoughts and emotions about the present, past, or future rather than taking action); emotional factors related to difficulties in identifying emotions, emotional unawareness, experiential avoidance, and feelings of meaninglessness (Mercer-Lynn, Hunter, & Eastwood, 2013); and physiological factors, which include non-optimal arousal and low levels of alertness (Hamilton, 1981).

Despite evidence highlighting the association between boredom and various negative mental health outcomes, research on boredom remains limited, necessitating further exploration across diverse experimental contexts (Koerth-Baker, 2016). To address this gap, several self-report assessments have been developed to measure states of boredom (for a review, see Mercer-Lynn et al., 2013). Multiple self-report scales have been created to characterize the neuropsychological signatures of boredom, each possessing a unique profile regarding the operationalization and measured aspects of this state. In this context, Fahlman et al. (2011) published the Multidimensional State Boredom Scale (MSBS), which is the only comprehensive measure of the state of boredom. The MSBS consists of 29 items that assess an individual’s experience of boredom in the moment. Participants respond to each item by indicating their level of agreement or disagreement using a 7-point Likert scale. The MSBS provides information about five factors/subscales: (a) disengagement; (b) high arousal negative affect; (c) low arousal negative affect; (d) inattention; and (e) time perception. This multidimensional nature of the MSBS offers valuable insights to deepen the understanding of the state of boredom in various psychopathological and psychosocial contexts.

### Objective

The MSBS could serve as a valuable tool for assessing boredom among individuals in Brazilian population studies. The aim of the present study is to translate the MSBS into Brazilian Portuguese and examine its psychometric properties. To achieve this, we sought to establish the psychometric properties of the Brazilian version of the MSBS by replicating the factor structure identified by Falhman et al. (2011) through confirmatory analysis.

## Methods

### Participants and Procedure

Data were collected from 329 volunteers (79% female and 21% male), with ages ranging from 12 to 68 years (M = 33.08, SD = 12.44). With permission from the original author of MSBS, the first author translated the original version of MSBS into Brazilian Portuguese. This version was verified through back-translation by a bilingual independent collaborator. The back-translated version was analyzed by Dr. John Eastwood, who was involved contributed to of the original version of the MSBS, to ensure reliability the translation. Thus, bilingual language experts and a boredom expert conducted translation and back-translation processes, faithful an accurate the MSBS into Brazilian Portuguese. The Brazilian version of the Multidimensional State Boredom Scale (MSBS) was evaluated by a committee of three experts, who assessed the items’ alignment with the purpose defined by the author of the original version and verify the semantic adequacy of the items in Portuguese. Subsequently, the preliminary version of the instrument was administered to 20 voluntaries aged raging 8-60 years, and they were asked about the clarity of comprehension of each item. No word needed to be rewritten (Beaton et al., 1998; Borsa et al., 2012).

To conduct an analysis of psychometric properties, participants were recruited through various online channels and social media communities in Brazil, including Facebook, WhatsApp, Telegram, and Instagram. This recruitment occurred via a link promoting a survey hosted on Google Forms over a five months period (november-april 2024). The research team shared the link, inviting individuals to participate voluntarily and anonymously, with no incentives were offered to participants. The eligibility criteria for the study required participants to be at least 12 years old and to speak Portuguese. The platform provided detailed information about the study’s objectives, and volunteers confirmed their participation by signing informed consent forms. On average, participants took 10 minutes to complete the preliminary version of the MSBS and a sociodemographic questionnaire. Participants provided informed consent for all experiments voluntarily, demonstrating their understanding and commitment to the study. This study was approved by the Federal University of Rio de Janeiro Ethics Committee (CAAE: 74921623.3.0000.5263), ensuring the ethical conduct of the research.

### Data Analysis

Confirmatory Factor Analysis (CFA) was conducted using the results from the adapted version to verify the factor structure model proposed by Fahlman et al. (2011). The model’s goodness-of-fit indices were evaluated using the Root Mean Square Error of Approximation (RMSEA), Comparative Fit Index (CFI), Tucker-Lewis Index (TLI), and Standardized Root Mean Square Residual (SRMR). The criteria suggested by DiStefano (2016) were employed as parameters to assess the model’s fit. Additionally, internal consistency was evaluated using Kaiser-Meyer-Olkin (KMO) test to verify the instrument’s reliability indicators, with a cut-off score above 0.7 considered acceptable for indicating reliability (Silva, 2019). The analysis utilized JASP version 0.18.3 for the data analysis.

## Results

### Confirmatory Factor Analysis (CFA)

The results of the CFA indicated a good factorial structure of the Brazilian version of MSBS: chi-square test (χ^2^) = 707.503, *df* = 367, *p* <0.001, with *χ*^2^/df = 1.92, and these results statistically rejecting the confirmatory factor model of the Portuguese version of the MSBS. Indeed, the root mean square error of approximation (RMSEA) was 0.079, greater than the conventional 0.05 value for a good model fit. The standardized root mean square residual (SRMSR) was 0.072, not close to the conventional 0.05 value for a good model fit. In addition, Bentler’s comparative fit index was 0.737, much lower than the required value of at least 0.90, leading us to conclude that the model was very poorly fitted. Therefore, considering that each of the four criteria above consistently testifies to an inadequate model fit, it is possible to conclude that the MSBS five-factor model proposed in the original validation study of the MSBS was adequate for this data. All factor loadings exhibited high and statistically significant values for all items (min = 0.43, max = 1.74; i.e., λij ≥ 0.50; see Table 1 for details). The item-total correlation was also satisfactory (min = 0.692, max = 0.924). Finally, the ECVI = 8.749.

**Table 1.**
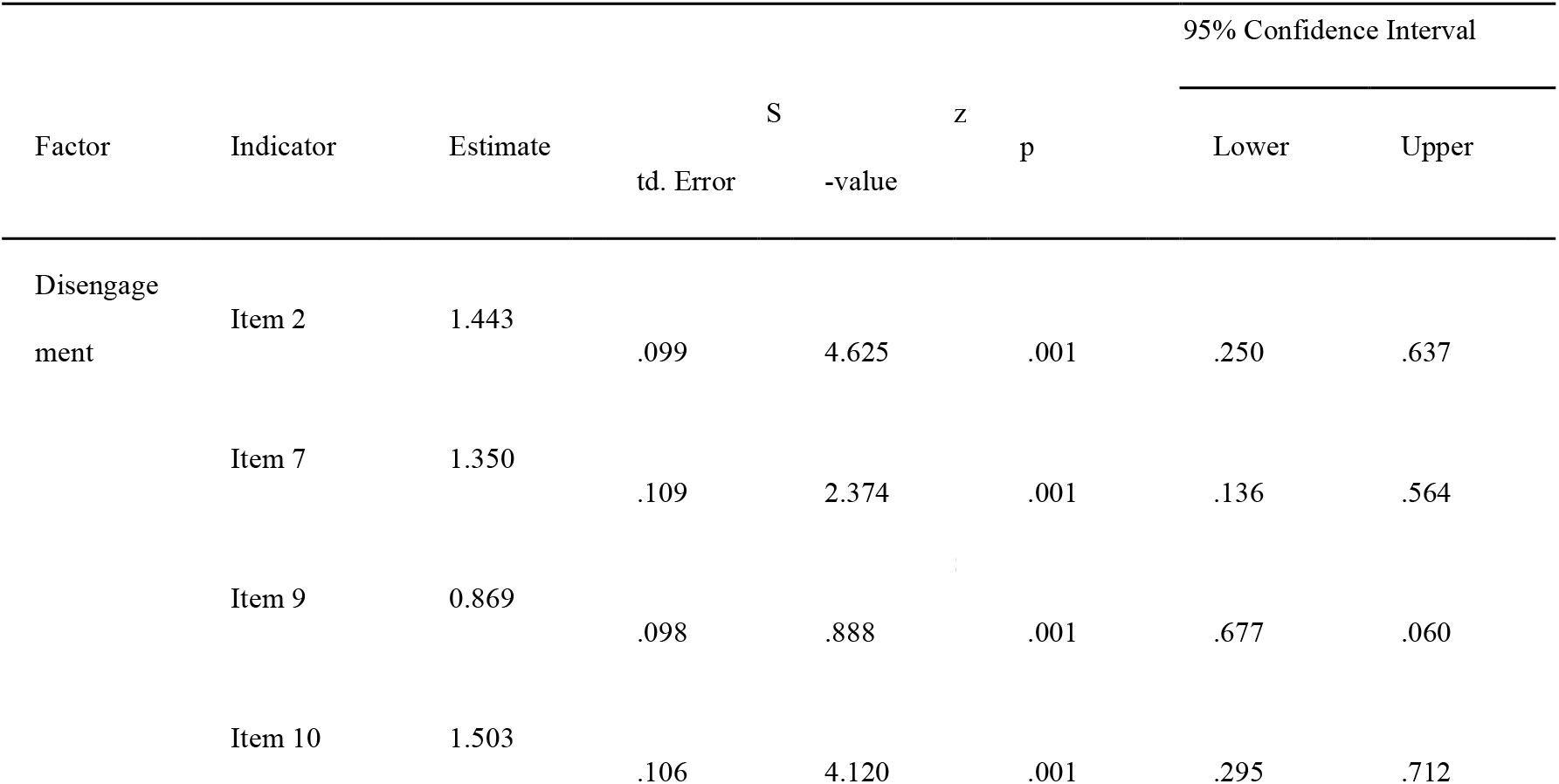
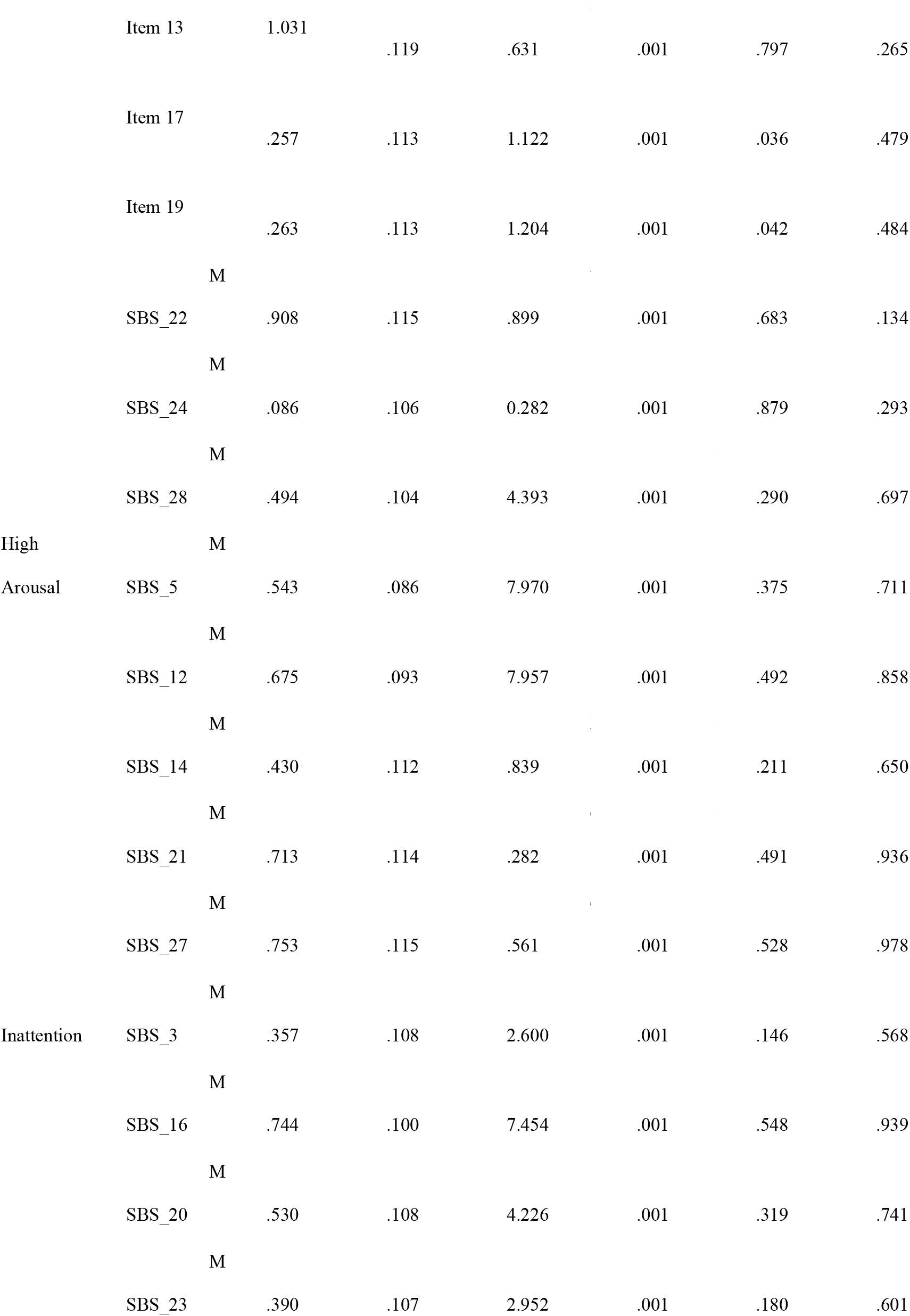
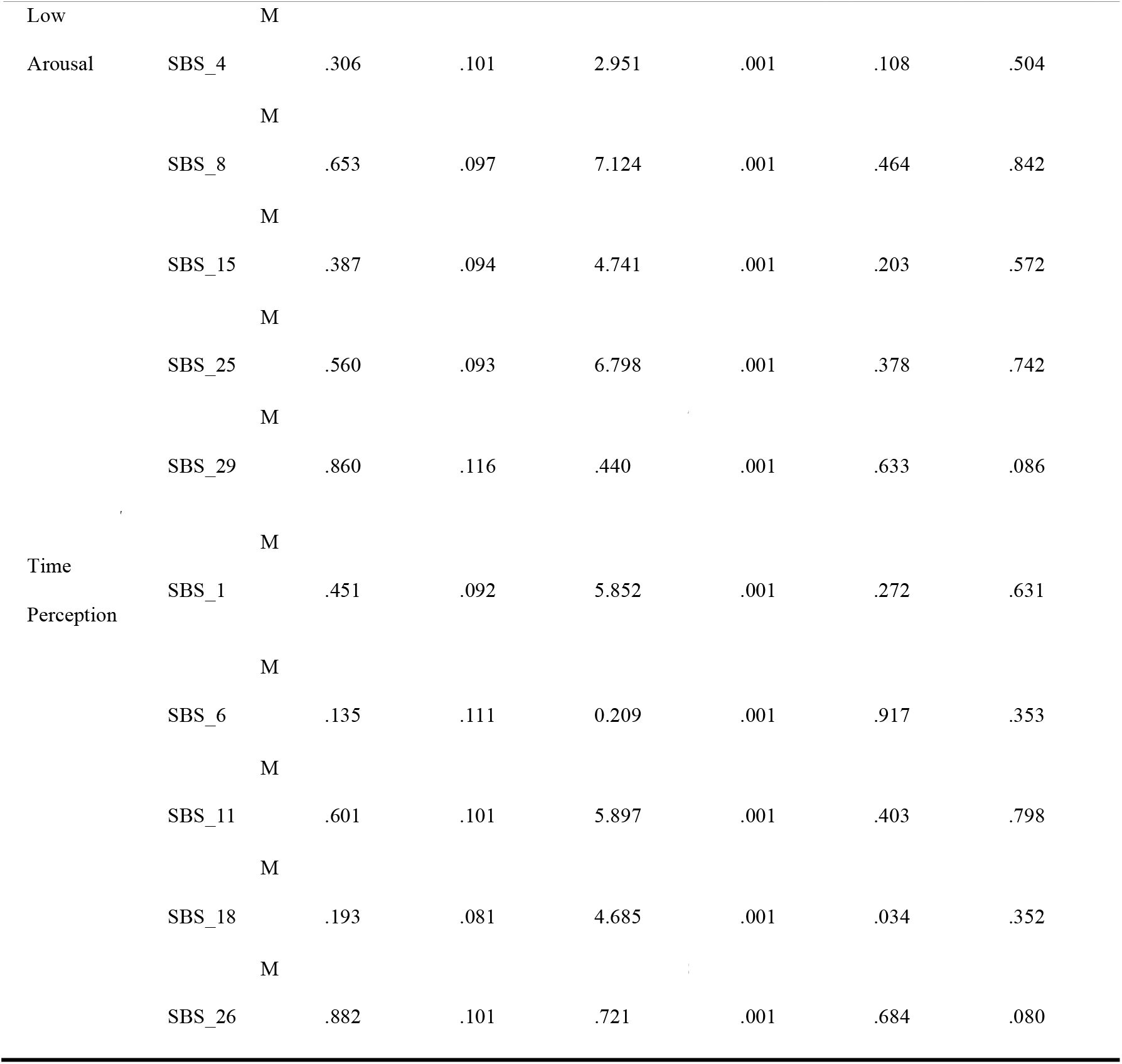
Factor loadings of items in the Brazilian version of MSBS.

### Internal Consistency

To test the reliability of the Brazilian version of MSBS, different reliability metrics (i.e., internal consistency), such as Cronbach’s alpha and McDonald’s omega, were used and analyzed. Cronbach’s alpha was 0.939 for total scores, 0.894 for Disengagement subscale; 0.781 for High Arousal subscale; 0.829 for Inattention subscale; 0.849 for Low Arousal subscale; and 0.861 for Time Perception subscale. There could not be improved by removing any items. Similarly, McDonald’s omega had a value of 0.939 for MSBS total scores; 0.891 for Disengagement subscale; 0.737 for High Arousal subscale; 0.825 for Inattention subscale; 0.840 for Low Arousal subscale; and 0.854 for Time Perception subscale. See the Table blow for details.

## Discussion

The Brazilian Portuguese version of the Multidimensional State Boredom Scale (MSBS) is considered a valid and reliable tool to assess the boredom state and exhibited the same factor structure as the original version showed in the study of Fahlman et al. (2013). Consistent with the original instrument, the relationships between factors remained moderate to strong; however, high arousal appeared to be less closely related to the other factors than what has been observed in the original version and in other studies of MSBS cross-cultural adaptation (Spoto et al., 2021).

The validation of MSBS could ensure the relevance and accuracy of the scale within a Brazilian context. If the validity of the MSBS is established in Brazil, it could yield significant benefits, including: (i) offering a reliable and concise tool for assessing the mental state of boredom in population-based studies, thereby addressing the current high demand for such assessments; (ii) equipping mental health promotion practitioners (e.g., physicians, psychologists) with a practical assessment tool for mental well-being, enabling them to evaluate the effectiveness of therapeutic strategies; and (iii) providing scientific researchers with the means to investigate the distribution and predictors of mental well-being, as well as various quality of life variables that can be disrupted by the experience of boredom.

The increasing popularity and utility of the MSBS across various cultural contexts have contributed to improving the scientific understanding of boredom as a mental phenomenon. Furthermore, comparative analyses of results from different countries can be useful in discovering whether the experience of boredom varies according to specific cultural factors, such as traditions and social engagement. Previous research suggests that Chinese participants reported lower levels of state boredom than North Americans when placed in similar circumstances. Authors propose that the experience of boredom can be influenced by distinct cultural traits, such as high positive arousal (Ng et al., 2015). Cultural aspects are significant in analyzing data collected from the Brazilian population, particularly due to the variability of internal and external factors. This understanding can help articulate the various ways in which cultural, cognitive, and psychopathological factors may impact the experience of boredom.

### Limitations and Future Directions

There are several limitations in the present study that can be addressed in future research. First, the current sample includes both adolescents and adults, resulting in a high level of participant heterogeneity. Therefore, it is important for the present findings to be replicated in larger samples and compared according to the developmental stages of the groups. Although other analyses did not show differences in the experience of boredom based on age, it will be relevant for future studies involving Brazilian samples to demonstrate the invariance of the MSBS factors across different age groups.

The study utilizing the original version of the MSBS demonstrates factor invariance based on gender (Fahlman et al., 2011). However, the present study enrolled a significantly higher number of females, and the sample size was insufficient to conduct a comparable gender-based factor invariance analysis of the Brazilian version of the MSBS. Additionally, this study did not account for the presence of psychopathology or various sociocultural factors (e.g., religion or educational levels) among the volunteers. Furthermore, the method of participant selection does not eliminate certain selection biases.

## Data Availability

All data produced in the present work are contained in the manuscript

## Acknowledgements

The author would like to express their sincere gratitude to professionals who contributed their time and effort to the translation step. We are also deeply thankful to Dr. John Eastwood, Ph.D., who provided invaluable support during all steps of this paper. Finally, we extend our appreciation to the reviewers and editors for their constructive feedback, which greatly improved the quality of this manuscript.

## Statements and Declarations

The author declare that he has no known competing financial interests or personal relationships that could have appeared to influence the work reported in this paper. Informed consent was obtained from all volunteers’ participants prior to their involvement in the study. This research did not receive any specific grant from funding agencies in the public, commercial, or not-for-profit sectors.

## References

Beaton, B., Bombardier, C., Guillemin, F., & Ferraz, M. (1998). Recommendations for the cross-cultural adaptation of health status measures. American Academy of Orthopaedic Surgeons Institute for Work & Health.

Biolcati, R., Passini, S., & Mancini, G. (2016). “I cannot stand the boredom.” Binge drinking expectancies in adolescence. Addictive Behaviors Reports, 3, 70–76. 10.1016/j.abrep.2016.05.001

Borsa, J. C., Damasio, B. F., & Bandeira, D. R. (2012). Adaptação e validação de instrumentos psicológicos entre culturas: Algumas considerações. Paidéia (Ribeirão Preto), 22(53), 123–130. 10.1590/S0103-863X2012000300014

Danckert, J., & Elpidorou, A. (2023). In search of boredom: Beyond a functional account. Trends in Cognitive Sciences, 27(6), 494–507. 10.1016/j.tics.2023.02.002

Eastwood, J. D., Frischen, A., Fenske, M. J., & Smilek, D. (2012). The unengaged mind: Defining boredom in terms of attention. Perspectives on Psychological Science, 7(5), 482– 495. 10.1177/1745691612456044

Fahlman, S. A., Mercer-Lynn, K. B., Flora, D. B., & Eastwood, J. D. (2013). Development and validation of the Multidimensional State Boredom Scale. Assessment, 20(1), 68–85. 10.1177/1073191111421303

Gerritsen, C. J., Toplak, M. E., Sciaraffa, J., & Eastwood, J. (2014). I can’t get no satisfaction: Potential causes of boredom. Consciousness and Cognition, 27, 27–41. 10.1016/j.concog.2013.10.001

Hamilton, J. A. (1981). Attention, personality, and the self-regulation of mood: Absorbing interest and boredom. Progress in Experimental Personality Research, 10, 281–315.

Hunter, A., & Eastwood, J. D. (2018). Does state boredom cause failures of attention? Examining the relations between trait boredom, state boredom, and sustained attention. Experimental Brain Research, 236(9), 2483–2492. 10.1007/s00221-016-4749-7

Isacescu, J., & Danckert, J. (2018). Exploring the relationship between boredom proneness and self-control in traumatic brain injury (TBI). Experimental Brain Research, 236(9), 2493– 2505. 10.1007/s00221-016-4682-9

Jarvis, W. B. G., & Seifert, T. F. (2002). Work avoidance as a function of task type and boredom proneness. Journal of Social Behavior and Personality, 17(1), 123–134.

Koerth-Baker, M. (2016). The bored mind is a hungry mind. The New York Times. https://www.nytimes.com

Lee, F. K. S., & Zelman, D. C. (2019). Boredom proneness as a predictor of depression, anxiety and stress: The moderating effects of dispositional mindfulness. Personality and Individual Differences, 146, 68– 75. 10.1016/j.paid.2019.04.001

Lipps, T. (1903). Leitfaden der Psychologie [Manual of Psychology]. Wilhelm Engelman Verlag.

Mercer-Lynn, K. B., Hunter, J. A., & Eastwood, J. D. (2013). Is trait boredom redundant? Journal of Social and Clinical Psychology, 32(8), 897–916. 10.1521/jscp.2013.32.8.897

Mikulas, W. L., & Vodanovich, S. J. (1993). The essence of boredom. Psychological Record, 43(1), 3–12.

Mugon, J., Struk, A., & Danckert, J. (2018). A failure to launch: The role of boredom in the emergence and persistence of failure to thrive. Personality and Individual Differences, 124, 1– 6. 10.1016/j.paid.2017.11.034

Ng, A. H., Liu, Y., Chen, J., & Eastwood, J. D. (2015). Culture and state boredom: A comparison between European Canadians and Chinese. Personality and Individual Differences, 75, 13– 18. 10.1016/j.paid.2014.10.053

Panda, P. K., et al. (2021). Psychological and behavioral impact of lockdown and quarantine measures for COVID-19 pandemic on children, adolescents and caregivers: A systematic review and meta-analysis. Journal of Tropical Pediatrics, 67(1), fmaa122. 10.1093/tropej/fmaa122

Spaeth, M., Weichold, K., & Silbereisen, R. K. (2015). The development of leisure boredom in early adolescence: Predictors and longitudinal associations with delinquency and depression. Developmental Psychology, 51(10), 1380. 10.1037/a0039480

Spoto, A., Iannattone, S., Valentini, P., Raffagnato, A., Miscioscia, M., & Gatta, M. (2021). Boredom in Adolescence: Validation of the Italian Version of the Multidimensional State Boredom Scale (MSBS) in Adolescents. Children (Basel, Switzerland), 8(4), 314. 10.3390/children8040314

Team J. (2020). JASP (Version 0.18.3) [Computer software]. https://jasp-stats.org/

